## Supplemental Information for "Regional connectivity drove bidirectional transmission of SARS-CoV-2 in the Middle East during travel restrictions"

### Supplementary information

#### Limited functional evidence for B.1.1.312 increased transmissibility suggests dominance due to epidemiological drivers

We observed a selective sweep of diversity to the B.1.1.312 lineage from August 2020 onwards, with the lineage accounting for 85% of all sampled cases after the putative onset of community transmission (**Main text Figure 2D**). We found that cases surged rapidly from September 2020 onwards, with the first wave peaking in November at 7933 cases and 91 deaths a day (**Main text Figure 1A**). It is difficult to accurately estimate when local transmission was first established with the temporal gaps in surveillance (**Main text Figure 1B**), but epidemiological evidence of the 10th death on the 3rd of July indicates a potential shift to community spread as the dominant source of new infections.

To investigate the timing and origin of the emergence of B.1.1.312, we reconstructed a lineage-specific phylogeographic build to ensure inclusion of all B.1.1.312 sequences worldwide (**Supplementary information figure 1A**). We estimated B.1.1.312 likely emerged in Jordan in late June [median tMRCA of 29 June, 95% HPD 7 June to 17 July], circulating cryptically for around 6 weeks owing to sparse sampling (**Supplementary information figure 1B**). We found that the B.1.1.312 sequences from Jordan formed a well-supported clade across all datasets, indicating that its growth did not result from multiple introduction events. We conclude that it either resulted from a single introduction that established onward transmission or that the lineage emerged locally in Jordan from a B.1.1 ancestor (**Supplementary information figure 1A**). We inferred the origin of the B.1.1.312 lineage as Jordan (posterior support 0.95). However, we estimated that the full B.1.1.312 lineage (including a basal USA sequence) emerged in late April [median 30 April, 95% HPD 1 April to 29 May], two months before sampled emergence in Jordan. This suggests we cannot exclude that the inferred local origin is driven by sampling rates of B.1.1.312 outside of Jordan, with our Jordanian sequences accounting for 87% of the lineage globally (**Supplementary information figure 1C**).

B.1.1.312's dominance during the first wave of infections raised concerns in Jordan about the possible increased intrinsic transmissibility of the lineage.[21,22] We estimated an exponential growth rate of 12.1 [95% HPD 9.12 - 15.12] for B.1.1.312 in our phylodynamic models. However, there were only 6 sequences generated from June to mid-August, limiting estimates on the relative growth rate of the lineage over time (**Figure 2B**).[71] Notably, B.1.1.312 carries the Q957L mutation in a conserved heptad repeat region (HRR) of the S2 subunit of the spike glycoprotein.[20] Interactions between the HRRs of the SARS-CoV-1 spike protein and HIV gp41 respectively were implicated in the conformational changes required for cell-cell fusion and viral entry, which raised concerns as to its phenotypic relevance.[20–22]

We characterized the phenotypic relevance of the Q957L substitution *in silico* and *in vitro* and found no functional advantage for Q957L, despite previous claims.[22] Changes at the 957 residue did not show a strong signal of convergent evolution in the global population and we did not find evidence that Q957L was under positive selection with the site-level Mixed Effects Model of Evolution (MEME) model across multiple datasets (**Supplementary information table 1**). Additionally, using the empirical force field algorithm FoldX, we estimated that Q957L has negligible stability effects on the spike protein ($\Delta\Delta G$ = -0.30 kcal/mol (S.D. = 0.03 kcal/mol), where $\Delta\Delta G$<-0.46 kcal/mol would be deemed to have stable effects by FoldX.

To validate the *in-silico* findings in an experimental context, we performed *in vitro* functional analyses on the Spike (S) protein with the Q957L mutation. We found that expression and processing of S was unaffected (**Supplementary information figure 2A**) and that infection of a lung cell line by lentiviral pseudotypes harboring the S constructs was not significantly different when bearing Q957L (**Supplementary information figure 2B**). Because the 957 residue is located within an HRR, we sought to further investigate a potential effect on S fusion activity. We used a cell-cell fusion assay in which effector cells expressing S WT or Q957L were co-cultured with cells expressing the viral receptor, the angiotensin converting enzyme 2 (Ace2), and an activating host serine protease, the transmembrane serine protease 2 (TMPRSS2). The effector and target cells are also co-transfected with fragments of the Venus fluorescent proteins fused with leucine zipper which can be reconstituted, and fluorescence measured when cell-cell fusion occurs. Using this assay, we did not detect a difference in the fusion activity of the S proteins (**Supplementary information figure 2C**), further suggesting no functional effect of the Q957L substitution.

B.1.1.312 emerged in a period of low incidence, which is associated with successful onward transmission of introductions (**Main text Figure 1A**).[7] Non-pharmaceutical interventions were completely relaxed during the estimated time of emergence (**Main text Figure 1A-C**). Eid Al-Adha was celebrated on July 31st in 2020, with holidays for public servants and many others observed around 30 July - 3 August. We observed an increase in mobility in this period compared to both the pre-pandemic baseline and the surrounding weeks across many geographic regions in Jordan, as many people traveled for the holidays (**Supplementary information figure 1D**). Notably, there was a 30+% increase in the first and third most populous cities of Amman and Zarqa on 31 July compared to baseline, which is 25% higher than the preceding days. The rapid surge in cases in September also coincided with more than two million students outside of designated high-incidence areas returning to in-person schooling in the first week (**Supplementary table 2**). Additionally. B.1.1.312 was also clearly displaced by the introduction of the B.1.1.7 Alpha variant in late November based on national prevalence estimates of SGTF and failed to establish itself after export to any regional or global country (**Supplementary figure 4)**. Taken together, we concluded that B.1.1.312 dominated the outbreak owing to stochastic, founder effects and epidemiological drivers during a period of low NPIs and limited population immunity as well as non-random sampling rather than intrinsic increased transmissibility.[8]

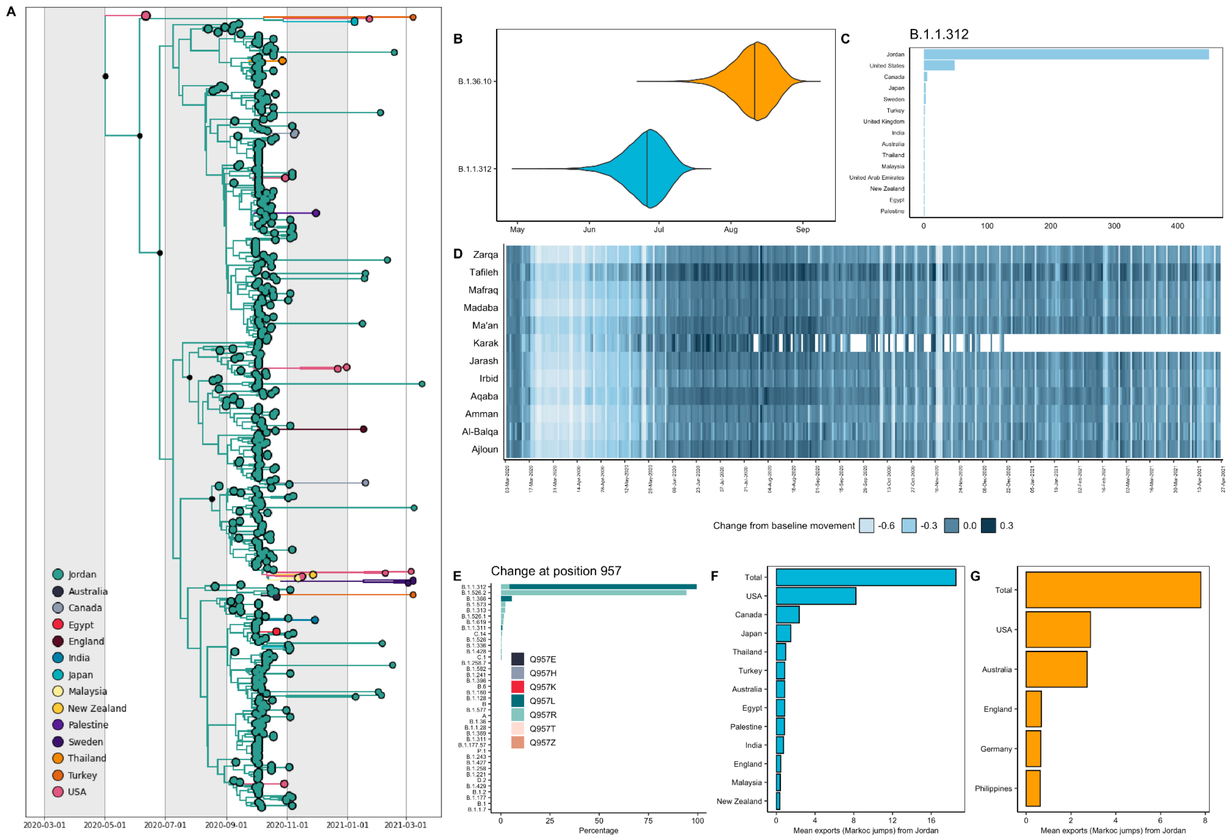


**Supplementary information figure 1: Characterization of B.1.1.312. A)** Time-calibrated phylogeny of the B.1.1.312 lineage. Branches are colored by country-level geographic state reconstruction. Internal nodes annotated with black point represent posterior support > 0.75.**B)** tMRCA distribution for the B.1.1.312 and B.1.36.10 lineages. **C)** Distribution of B.1.1.312 outside of Jordan up to October 2021

**D)** Facebook change in movement data for all major governorates. **E)** Change at residue 957 across all lineages with variability at the position **F)** Posterior mean number of exports from Jordan into each source country for the B.1.1.312 lineage for all statistically supported transitions (BF>3). **G)** Mean markov jumps from Jordan into each source country for the B.1.36.10 lineage for all statistically supported transitions (BF>3).

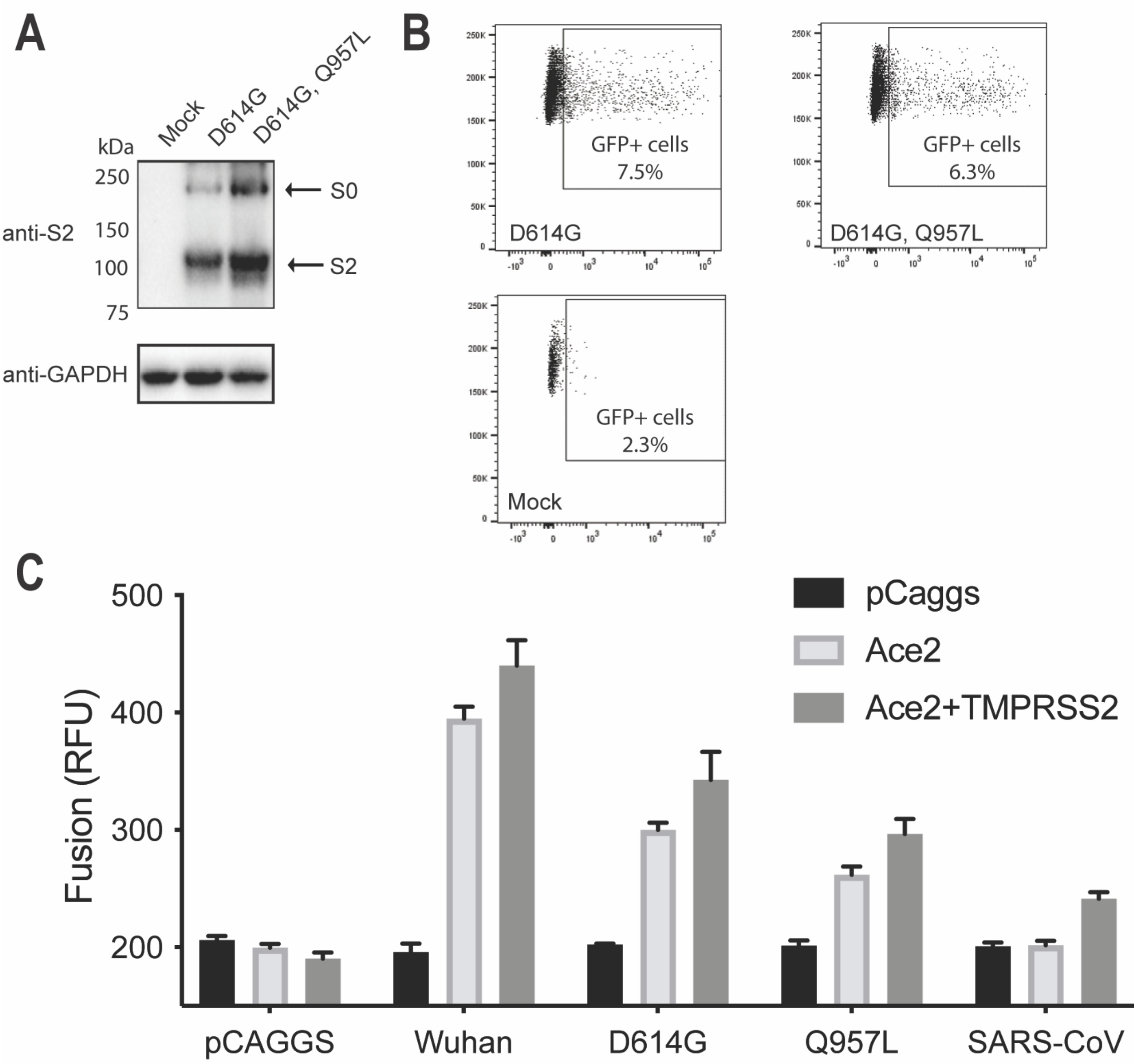


**Supplemental information figure 2. Functional characterization of SARS-CoV-2 Spike carrying the Q957L mutation.** HEK 293T cells were transfected to produce GFP encoding lentiviral pseudotypes bearing the SARS-CoV-2 Spike (S) D614G, D614G, Q957L mutations, or no S (Mock). (A) Cell lysates were resolved by SDS-PAGE and GAPDH, S0 and processed S2 were detected by immunoblot. (B) Equivalent amounts of supernatant were used to infect Calu-3 cells. GFP positive cells were quantified by flow cytometry 48 hours post-infection (Representative experiments of n=3 is shown). The impact of the Q957L mutation on S fusion activity was directly assessed by bi-molecular fluorescence complementation using effector HEK 293T cells expressing ZipV1 with or without the S constructs and target HEK 293T cells expressing ZipV2, Ace2 in the presence or not of TMPRSS2. Fusion was measured by detection of reconstituted Venus fluorescence. (Data are average of 3 independent experiments)

**Supplementary information table 1**: MEME results (see https://github.com/andersen-lab/paper_2022_jordan-sars2-phylogenetics)

| Site | alpha | beta | p<sup>-</sup> | β<sup>+</sup> | p<sup>+</sup> | LRT | p-value | # branches under selection | Total branch length | MEME LogL | FEL LogL | Replicate |
|---|---|---|---|---|---|---|---|---|---|---|---|---|
| 33 | 0 | 0 | 1 | 1360.48 | 0 | 10.6 | 0 | 1 | 0 | -18.05 | -13.11 | 8 |
| 70 | 0 | 0 | 0.01 | 4.49 | 0.99 | 3.06 | 0.1 | 3 | 0 | -25.05 | -25.05 | 8 |
| 138 | 0 | 0 | 0.52 | 27.55 | 0.48 | 3.24 | 0.09 | 7 | 0 | -51.69 | -51.68 | 8 |
| 262 | 0 | 0 | 0.01 | 7.99 | 0.99 | 3.06 | 0.1 | 5 | 0 | -36.74 | -36.74 | 8 |
| 291 | 0 | 0 | 1 | 2035.88 | 0 | 12.25 | 0 | 1 | 0 | -19.57 | -13.78 | 8 |
| 439 | 0 | 0 | 0.5 | 19.93 | 0.5 | 5 | 0.04 | 3 | 0 | -30.33 | -30.33 | 8 |
| 452 | 0 | 0 | 0.01 | 9.71 | 0.99 | 7.85 | 0.01 | 4 | 0 | -31.44 | -31.45 | 8 |
| 501 | 0 | 0 | 0.95 | 373.62 | 0.05 | 4.76 | 0.04 | 5 | 0 | -43.12 | -42.45 | 8 |
| 614 | 0 | 0 | 0 | 25.84 | 1 | 10.21 | 0 | 18 | 0 | -116.36 | -116.36 | 8 |
| 675 | 0 | 0 | 0 | 17.57 | 1 | 3.83 | 0.07 | 9 | 0 | -58.13 | -58.13 | 8 |
| 677 | 0 | 0 | 0 | 17.94 | 1 | 4.01 | 0.06 | 9 | 0 | -57.66 | -57.66 | 8 |
| 681 | 0 | 0 | 0.05 | 8.02 | 0.95 | 3.07 | 0.1 | 5 | 0 | -46.3 | -46.3 | 8 |
| 1118 | 0 | 0 | 0.53 | 17.32 | 0.47 | 4.85 | 0.04 | 4 | 0 | -35.02 | -35 | 8 |
| 1167 | 0 | 0 | 0.98 | 219.85 | 0.02 | 7.84 | 0.01 | 3 | 0 | -29.67 | -26.9 | 8 |
| 5 | 0 | 0 | 0 | 21.88 | 1 | 10.79 | 0 | 18 | 0 | -105.7 | -105.7 | 1 |
| 22 | 0 | 0 | 0.01 | 8.48 | 0.99 | 4 | 0.06 | 5 | 0 | -35.55 | -35.55 | 1 |
| 26 | 0 | 0 | 0.01 | 6.87 | 0.99 | 3.44 | 0.08 | 6 | 0 | -42.49 | -42.49 | 1 |
| 95 | 0 | 0 | 0.01 | 8.34 | 0.99 | 3.99 | 0.06 | 5 | 0 | -33.87 | -33.87 | 1 |
| 98 | 0 | 0 | 0.05 | 8.48 | 0.95 | 4.09 | 0.06 | 7 | 0 | -49.3 | -49.3 | 1 |
| 178 | 0 | 0 | 0.54 | 10.72 | 0.46 | 3.79 | 0.07 | 3 | 0 | -29.42 | -29.41 | 1 |
| 452 | 0 | 0 | 0.01 | 10.99 | 0.99 | 11.23 | 0 | 6 | 0 | -44.1 | -44.1 | 1 |
| 477 | 0 | 0 | 0.01 | 9.16 | 0.99 | 7.07 | 0.01 | 6 | 0 | -46.43 | -46.43 | 1 |
| 501 | 0 | 0 | 0.87 | 167.57 | 0.13 | 8.04 | 0.01 | 8 | 0 | -60.93 | -60.33 | 1 |
| 677 | 0 | 0 | 0 | 18.64 | 1 | 4.39 | 0.05 | 11 | 0 | -74.9 | -74.9 | 1 |
| 681 | 0 | 0 | 0 | 20.87 | 1 | 10.41 | 0 | 18 | 0 | -123.59 | -123.59 | 1 |
| 813 | 0 | 0 | 0.04 | 4.61 | 0.96 | 3.49 | 0.08 | 3 | 0 | -28.08 | -28.08 | 1 |
| 1078 | 0 | 0 | 0 | 8.46 | 1 | 4.27 | 0.05 | 7 | 0 | -48.06 | -48.01 | 1 |
| 5 | 0 | 0 | 0 | 16.47 | 1 | 8.55 | 0.01 | 13 | 0 | -87.94 | -87.94 | 2 |
| 18 | 0 | 0 | 0 | 10.65 | 1 | 5.64 | 0.03 | 10 | 0 | -62.09 | -62.09 | 2 |
| 95 | 0 | 0 | 0.01 | 8.28 | 0.99 | 4.12 | 0.06 | 5 | 0 | -32.09 | -32.09 | 2 |
| 222 | 0 | 0 | 0.01 | 9.47 | 0.99 | 4.98 | 0.04 | 8 | 0 | -53.9 | -53.9 | 2 |
| 262 | 0 | 0 | 0.53 | 13.16 | 0.47 | 3.14 | 0.1 | 5 | 0 | -40.65 | -40.64 | 2 |
| 367 | 0 | 0 | 0.01 | 3.54 | 0.99 | 3.34 | 0.09 | 3 | 0 | -24.53 | -24.53 | 2 |
| 501 | 0 | 0 | 0.93 | 318.59 | 0.07 | 7.24 | 0.01 | 7 | 0 | -55.62 | -54.8 | 2 |
| 505 | 0 | 0 | 0.99 | 468.19 | 0.01 | 9.85 | 0 | 1 | 0 | -20.7 | -17.1 | 2 |
| 614 | 0 | 0 | 0.01 | 6.04 | 0.99 | 3.17 | 0.1 | 5 | 0 | -41.86 | -41.86 | 2 |
| 675 | 0 | 0 | 0 | 21.01 | 1 | 4.68 | 0.04 | 14 | 0 | -88.02 | -88.02 | 2 |
| 677 | 0 | 0 | 0 | 25.29 | 1 | 5.18 | 0.03 | 16 | 0 | -93.63 | -93.62 | 2 |
| 679 | 0 | 0 | 0.52 | 21.09 | 0.48 | 3.08 | 0.1 | 4 | 0 | -39.06 | -39.05 | 2 |
| 681 | 0 | 0 | 0.01 | 13.27 | 0.99 | 6.97 | 0.01 | 12 | 0 | -93.58 | -93.58 | 2 |
| 939 | 0 | 0 | 0.01 | 6.59 | 0.99 | 3.52 | 0.08 | 6 | 0 | -42.56 | -42.56 | 2 |
| 1176 | 0 | 0 | 0.01 | 8.14 | 0.99 | 4.2 | 0.06 | 7 | 0 | -49.65 | -49.65 | 2 |
| 5 | 0 | 0 | 0 | 20.54 | 1 | 10.2 | 0 | 17 | 0 | -102.87 | -102.87 | 3 |
| 18 | 0 | 0 | 0 | 8 | 1 | 3.94 | 0.07 | 7 | 0 | -46.27 | -46.27 | 3 |
| 76 | 0 | 0 | 0.04 | 8.99 | 0.96 | 4 | 0.06 | 5 | 0 | -36.49 | -36.49 | 3 |
| 95 | 0 | 0 | 0.01 | 6.87 | 0.99 | 3.19 | 0.1 | 4 | 0 | -27.85 | -27.85 | 3 |
| 157 | 0 | 0 | 0.01 | 4.34 | 0.99 | 3.48 | 0.08 | 4 | 0 | -32.01 | -32.01 | 3 |
| 316 | 1.01 | 0 | 1 | 10000 | 0 | 19.84 | 0 | 1 | 0 | -25.93 | -16.06 | 3 |
| 367 | 0 | 0 | 0.01 | 3.85 | 0.99 | 3.3 | 0.09 | 3 | 0 | -26.1 | -26.1 | 3 |
| 439 | 0 | 0 | 0.5 | 21.09 | 0.5 | 6.65 | 0.02 | 4 | 0 | -37.91 | -37.91 | 3 |
| 452 | 0 | 0 | 0 | 12.63 | 1 | 12.78 | 0 | 7 | 0 | -49.62 | -49.62 | 3 |
| 501 | 0 | 0 | 0.86 | 218.67 | 0.14 | 9.45 | 0 | 11 | 0 | -80.8 | -79.78 | 3 |
| 655 | 0 | 0 | 0 | 10.07 | 1 | 3.13 | 0.1 | 7 | 0 | -43.05 | -43.05 | 3 |
| 677 | 0 | 0 | 0 | 26.86 | 1 | 5.73 | 0.03 | 16 | 0 | -100.11 | -100.11 | 3 |
| 681 | 0 | 0 | 0 | 16.12 | 1 | 8 | 0.01 | 14 | 0 | -103.25 | -103.26 | 3 |
| 943 | 0 | 0 | 0.99 | 2851.3 | 0.01 | 32.1 | 0 | 3 | 0 | -48.3 | -33.61 | 3 |
| 95 | 0 | 0 | 0.01 | 15.68 | 0.99 | 5.83 | 0.02 | 6 | 0 | -45.37 | -45.37 | 7 |
| 178 | 0 | 0 | 0.54 | 13.22 | 0.46 | 3.6 | 0.08 | 3 | 0 | -28.72 | -28.71 | 7 |
| 222 | 0 | 0 | 0 | 7.59 | 1 | 3.09 | 0.1 | 5 | 0 | -34.49 | -34.49 | 7 |

| | | | | | | | | | | | | |
|---|---|---|---|---|---|---|---|---|---|---|---|---|
| 439 | 0 | 0 | 0.54 | 14.83 | 0.46 | 3.42 | 0.09 | 2 | 0 | -21.74 | -21.74 | 7 |
| 452 | 2.87 | 0.1 | 0 | 11.47 | 1 | 3.07 | 0.1 | 5 | 0 | -54.9 | -54.9 | 7 |
| 477 | 0 | 0 | 0.01 | 7.63 | 0.99 | 4.39 | 0.05 | 4 | 0 | -33.02 | -33.02 | 7 |
| 501 | 0 | 0 | 0.96 | 575 | 0.04 | 4.61 | 0.05 | 5 | 0 | -43.07 | -42.52 | 7 |
| 614 | 0 | 0 | 0 | 12.27 | 1 | 4.69 | 0.04 | 8 | 0 | -59.62 | -59.62 | 7 |
| 615 | 0 | 0 | 0.99 | 191.47 | 0.01 | 6.02 | 0.02 | 1 | 0 | -16.87 | -13.86 | 7 |
| 677 | 0 | 0 | 0.01 | 22.58 | 0.99 | 4.11 | 0.06 | 12 | 0 | -72.18 | -72.18 | 7 |
| 681 | 0 | 0 | 0.52 | 22.45 | 0.48 | 4.3 | 0.05 | 7 | 0 | -57.89 | -57.87 | 7 |
| 845 | 0 | 0 | 0.02 | 7.89 | 0.98 | 3.1 | 0.1 | 5 | 0 | -38.06 | -38.06 | 7 |
| 879 | 0 | 0 | 0.01 | 4.6 | 0.99 | 3.12 | 0.1 | 3 | 0 | -26.1 | -26.1 | 7 |
| 936 | 0 | 0 | 0.53 | 17.39 | 0.47 | 4.79 | 0.04 | 4 | 0 | -33.01 | -33 | 7 |
| 1058 | 8.11 | 0 | 0.99 | 177.57 | 0.01 | 3.4 | 0.09 | 1 | 0 | -22.54 | -19.47 | 7 |
| 26 | 0 | 0 | 0.99 | 629.98 | 0.01 | 9.84 | 0 | 2 | 0 | -24.85 | -20.77 | 6 |
| 95 | 0 | 0 | 0.01 | 9 | 0.99 | 3.18 | 0.1 | 4 | 0 | -26.93 | -26.93 | 6 |
| 439 | 0 | 0 | 0.54 | 15.9 | 0.46 | 3.51 | 0.08 | 2 | 0 | -21.28 | -21.28 | 6 |
| 452 | 1.64 | 0.08 | 0.01 | 12.58 | 0.99 | 4.86 | 0.04 | 5 | 0 | -46.5 | -46.5 | 6 |
| 477 | 0 | 0 | 0.01 | 8.41 | 0.99 | 4.65 | 0.05 | 4 | 0 | -36.62 | -36.62 | 6 |
| 501 | 0 | 0 | 0.94 | 425.98 | 0.06 | 5.84 | 0.02 | 6 | 0 | -49.07 | -48.46 | 6 |
| 572 | 0 | 0 | 0.01 | 9.08 | 0.99 | 3.19 | 0.1 | 4 | 0 | -27.72 | -27.72 | 6 |
| 614 | 0.02 | 0.01 | 0.51 | 20.32 | 0.49 | 3.65 | 0.08 | 6 | 0 | -48.25 | -48.24 | 6 |
| 677 | 0.01 | 0 | 0 | 28.77 | 1 | 4.48 | 0.05 | 14 | 0 | -80.43 | -79.89 | 6 |
| 5 | 0 | 0 | 0 | 16.83 | 1 | 7.86 | 0.01 | 14 | 0 | -85.77 | -85.77 | 4 |
| 18 | 0 | 0 | 0 | 10.78 | 1 | 5.03 | 0.04 | 9 | 0 | -55.84 | -55.84 | 4 |
| 95 | 0 | 0 | 0 | 12.35 | 1 | 5.49 | 0.03 | 7 | 0 | -44.76 | -44.76 | 4 |
| 138 | 0 | 0 | 0 | 18.59 | 1 | 5.53 | 0.03 | 11 | 0 | -75.54 | -75.54 | 4 |
| 222 | 0 | 0 | 0.04 | 7.94 | 0.96 | 3.64 | 0.08 | 6 | 0 | -43.12 | -43.12 | 4 |
| 439 | 0 | 0 | 0.53 | 11.46 | 0.47 | 3.35 | 0.09 | 2 | 0 | -21.92 | -21.91 | 4 |
| 452 | 1.27 | 0.1 | 0 | 16.81 | 1 | 10.93 | 0 | 9 | 0 | -68.21 | -68.21 | 4 |
| 501 | 0 | 0 | 0.93 | 436.6 | 0.07 | 10.24 | 0 | 8 | 0 | -69.78 | -68.04 | 4 |
| 512 | 0 | 0 | 0.99 | 161.79 | 0.01 | 4.76 | 0.04 | 1 | 0 | -17.34 | -14.88 | 4 |
| 614 | 0 | 0 | 0 | 6.47 | 1 | 3.08 | 0.1 | 5 | 0 | -40.37 | -40.37 | 4 |
| 677 | 0 | 0 | 0 | 26.67 | 1 | 5.65 | 0.03 | 17 | 0 | -99.35 | -99.16 | 4 |
| 681 | 0 | 0 | 0.02 | 12.05 | 0.98 | 5.6 | 0.03 | 10 | 0 | -78.55 | -78.55 | 4 |
| 816 | 0 | 0 | 1 | 528.82 | 0 | 9.45 | 0 | 1 | 0 | -18.67 | -14.21 | 4 |
| 859 | 0 | 0 | 0.01 | 8.7 | 0.99 | 3.94 | 0.07 | 5 | 0 | -35.9 | -35.9 | 4 |
| 1176 | 0 | 0 | 0.24 | 10.16 | 0.76 | 3.61 | 0.08 | 6 | 0 | -45.68 | -45.49 | 4 |
| 5 | 0 | 0 | 0 | 22.88 | 1 | 12.17 | 0 | 21 | 0 | -115.61 | -115.61 | 5 |
| 18 | 0 | 0 | 0 | 8.56 | 1 | 4.58 | 0.05 | 7 | 0 | -53.66 | -53.66 | 5 |
| 67 | 0 | 0 | 0.01 | 7.07 | 0.99 | 3.78 | 0.07 | 6 | 0 | -43.82 | -43.82 | 5 |
| 95 | 0 | 0 | 0 | 11.68 | 1 | 5.8 | 0.02 | 7 | 0 | -42.63 | -42.63 | 5 |
| 98 | 0 | 0 | 0.53 | 11.43 | 0.47 | 3.17 | 0.1 | 5 | 0 | -39.51 | -39.5 | 5 |
| 222 | 0 | 0 | 0.01 | 10.74 | 0.99 | 5.67 | 0.03 | 9 | 0 | -58.8 | -58.8 | 5 |
| 439 | 0 | 0 | 0.54 | 17.24 | 0.46 | 5.35 | 0.03 | 3 | 0 | -29.05 | -29.05 | 5 |
| 452 | 0 | 0 | 0.01 | 12.32 | 0.99 | 13.15 | 0 | 7 | 0 | -50.92 | -50.93 | 5 |
| 501 | 0 | 0 | 0.96 | 413.24 | 0.04 | 7.13 | 0.01 | 6 | 0 | -50.32 | -49.07 | 5 |
| 572 | 0 | 0 | 0.01 | 9.86 | 0.99 | 4.99 | 0.04 | 6 | 0 | -41.44 | -41.44 | 5 |
| 614 | 0 | 0 | 0.08 | 7.98 | 0.92 | 3.85 | 0.07 | 6 | 0 | -50.52 | -50.48 | 5 |
| 675 | 0 | 0 | 0 | 14.88 | 1 | 3.21 | 0.1 | 10 | 0 | -68.87 | -68.87 | 5 |
| 677 | 0 | 0 | 0 | 25.03 | 1 | 5.24 | 0.03 | 17 | 0 | -100.68 | -100.68 | 5 |
| 681 | 0 | 0 | 0.01 | 21.6 | 0.99 | 11.3 | 0 | 18 | 0 | -136.23 | -136.23 | 5 |
| 780 | 0.08 | 0.03 | 0.99 | 1715.22 | 0.01 | 10.51 | 0 | 2 | 0 | -27.3 | -22.42 | 5 |
| 859 | 0 | 0 | 0.01 | 11.5 | 0.99 | 5.82 | 0.02 | 7 | 0 | -46.79 | -46.79 | 5 |
| 943 | 0 | 0 | 1 | 2822.01 | 0 | 14.64 | 0 | 2 | 0 | -29.49 | -22.89 | 5 |

### Supplementary Figures from main text

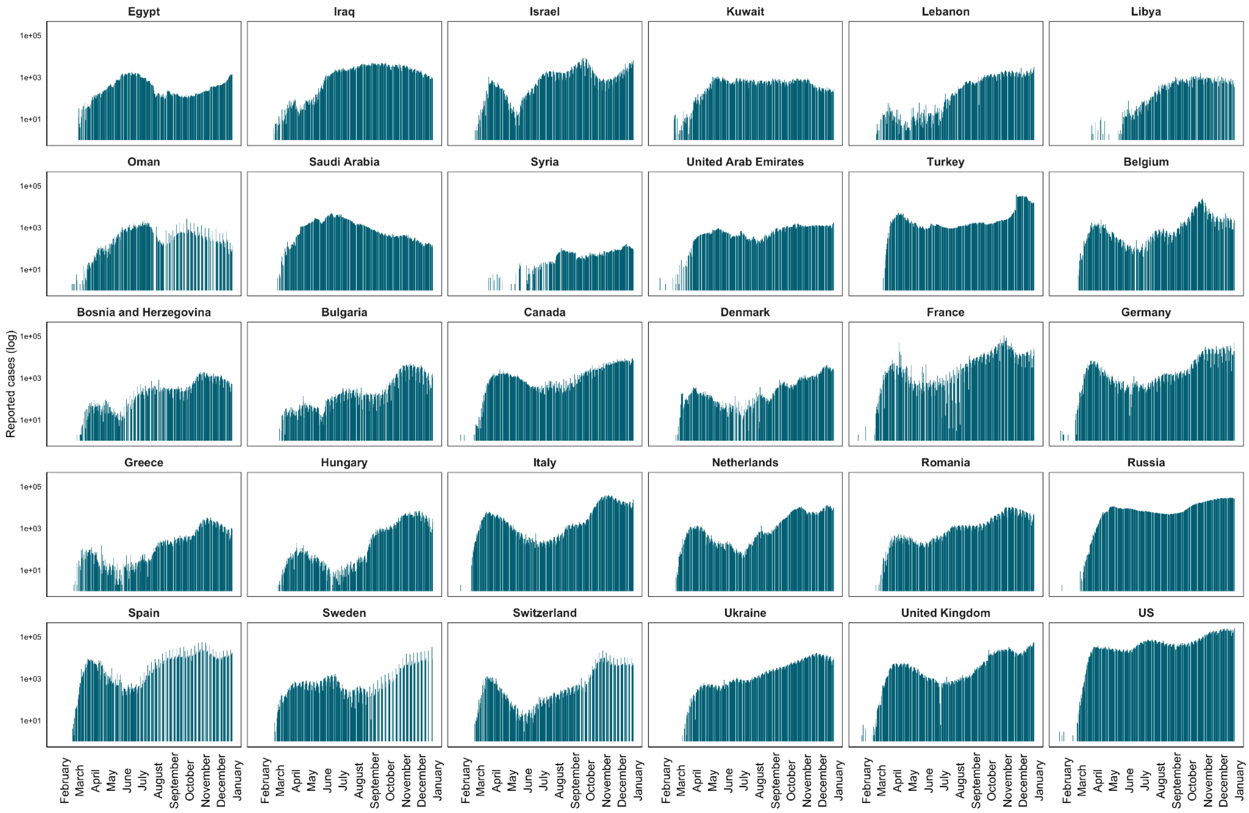


Supplementary figure 1: Log-scale daily cases for top contributing countries to III (including all Middle Eastern countries)

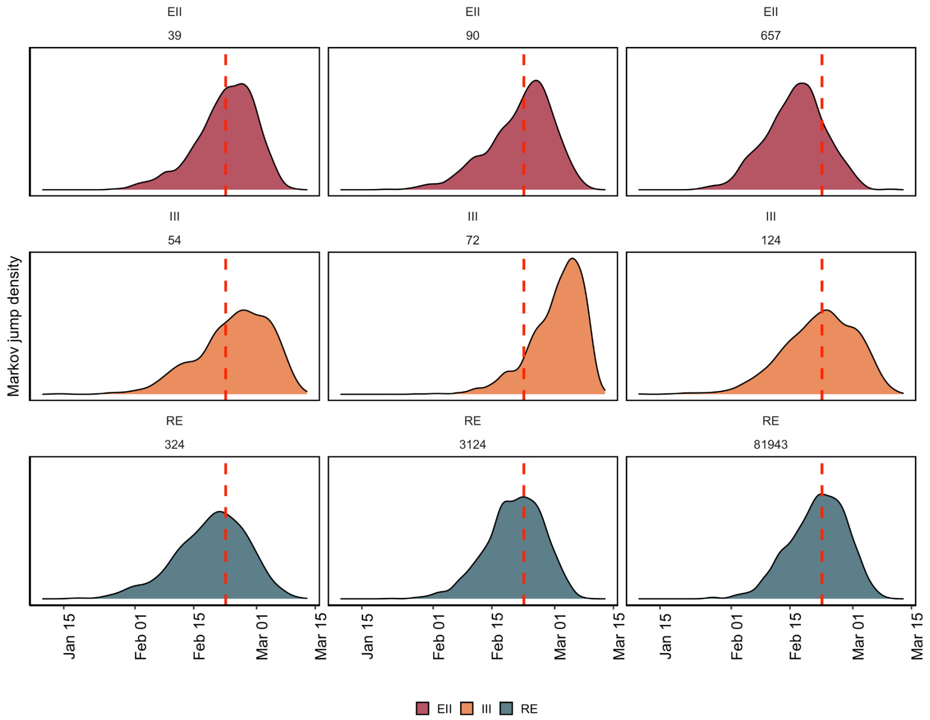


Supplementary figure 2: Markov jump density for the initial introduction into Jordan, summarized across all global datasets. Dashed line indicates the mean.

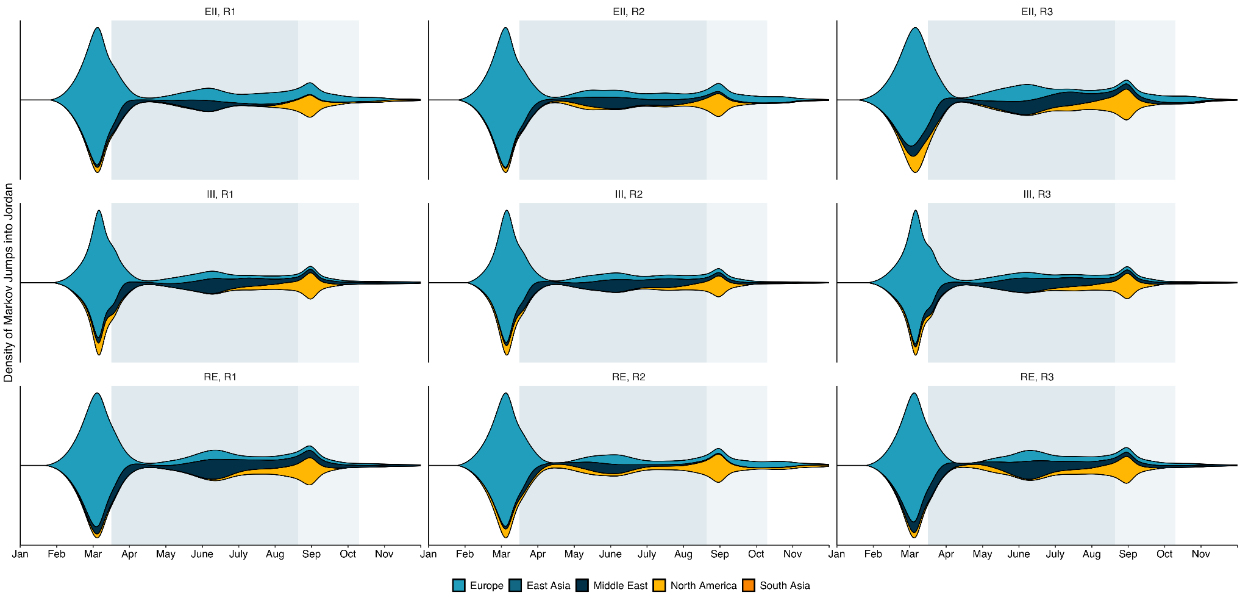


Supplementary figure 3: The density of estimated introductions (Markov jumps) into Jordan over time from source regions by downsampling strategy (EII, III, RE) for the three replicates (indicated by Rx in title). Period of full travel restrictions (March - September) highlighted in darker blue, with extended land border closure up to the end of October highlighted in lighter blue. EII: Epidemiological incidence informed; RE: Regional enrichment; III: Introduction Intensity Index informed

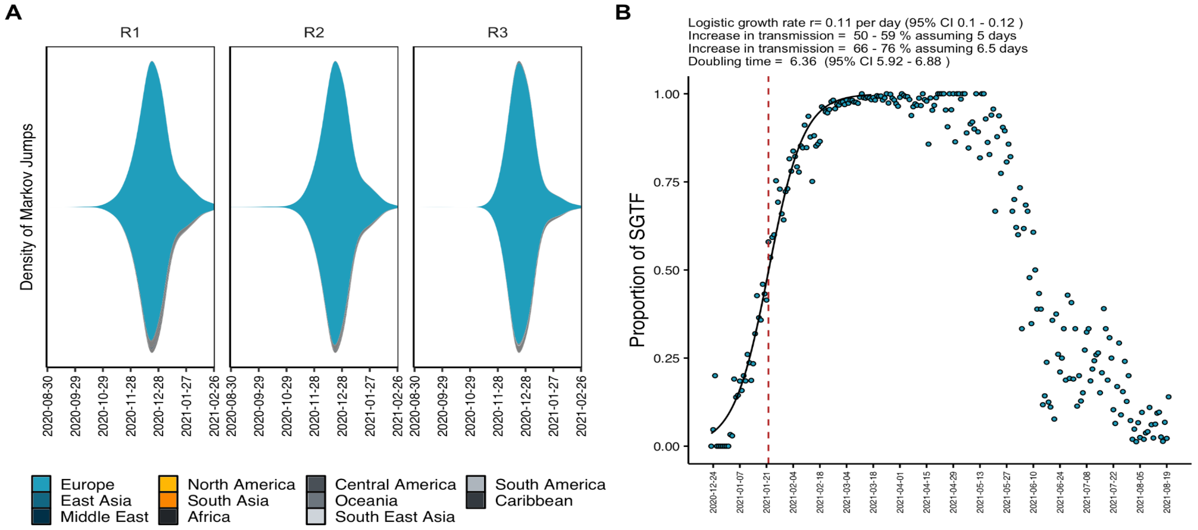


Supplementary figure 4: A) The density of estimated introductions (Markov jumps) into Jordan for the Alpha variant (B.1.1.7) over time from source regions for random replicates (indicated by R1-3) B) Logistic growth model fit to proportion of SGTF in total positive tests performed at Biolab Diagnostic Laboratories in Jordan over the initial 3-month period. Red line indicates the number of days since first detection when the proportion of B.1.1.7 cases crossed 50%.

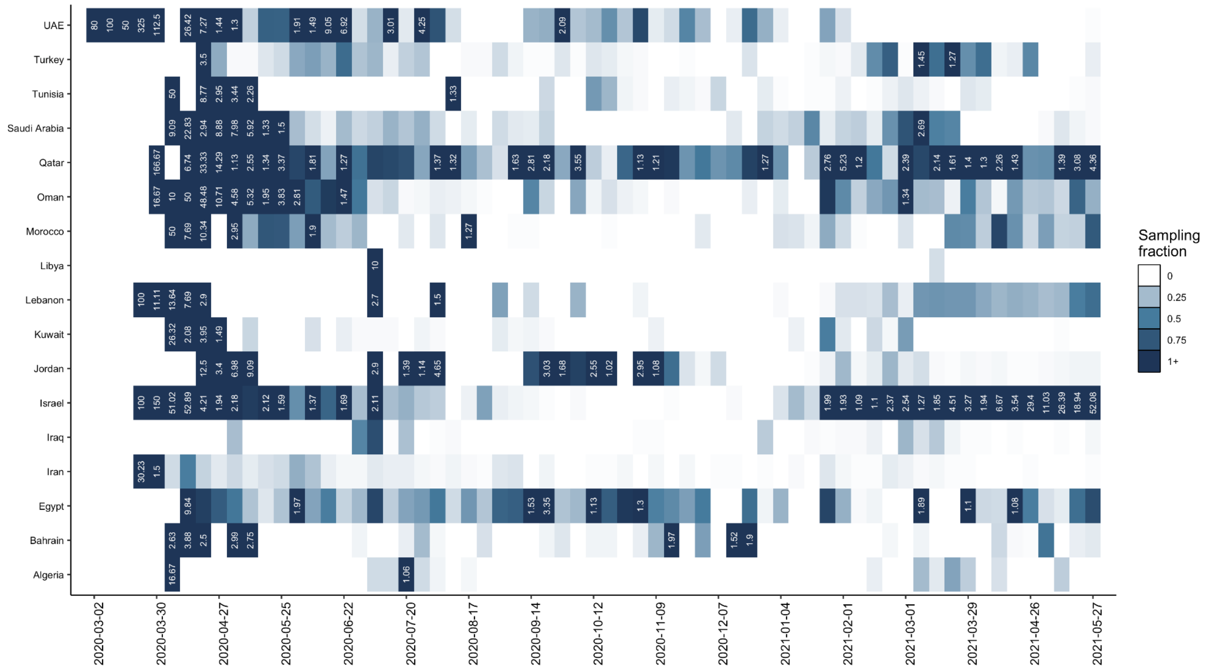


Supplementary figure 5: Dissimilarity between weekly binned lineage frequency profiles as presence-absence of lineages between Jordan and its MENA neighbors by Bray-Curtis dissimilarity. Bray-Curtis dissimilarity is bound between 0 and 1, with 0 indicating the countries have the same composition and 1 indicates countries do not share any lineages. **H)** Weekly-rolling average sampling fraction for MENA countries. All weeks with a sampling fraction above 1% are annotated with their sampling fraction in text for visualization scale.

Supplementary table 1: Posterior support for transitions into and out of Jordan in Markov jump estimation. Only transition rates with a Bayes factor higher than 3 are reported.

Supplementary table 2: Timeline of non-pharmaceutical interventions in Jordan (see https://github.com/andersen-lab/paper_2022_jordan-sars2-phylogenetics)

Supplementary table 3: GISAID acknowledgements Jordan (see https://github.com/andersen-lab/paper_2022_jordan-sars2-phylogenetics)
